# Genetic risk impacts stroke mortality and pathogenesis in patients with ischemic stroke: cohort study of BioBank Japan

**DOI:** 10.1101/2024.12.08.24318693

**Authors:** Takashi Shimoyama, Yoichiro Kamatani, Koichi Matsuda, Hiroki Yamaguchi, Kazumi Kimura

## Abstract

**Background:** Previous multi-ancestry genome-wide association studies (GWAS) of stroke reported 32 stroke risk loci in the MEGASTROKE study. Most studies on the genetic risk score (GRS) of stroke have reported a predominance in the European general population. We aimed to explore the association among GRS, clinical characteristics, and mortality in patients with ischemic stroke registered in the BioBank Japan (BBJ) database.

**Methods:** This is a cohort study of BBJ participants. The project participants were recruited between June 2003 and March 2018. We conducted a GWAS for stroke in 19,702 Japanese patients with ischemic stroke (controls, n=159,610). GRS was generated using 29 stroke risk single nucleotide polymorphisms (SNPs) from 32 stroke-related loci identified in the MEGASTROKE. A multivariate logistic regression model was used to estimate odds ratios (ORs) and 95% confidence intervals (95% CIs) for comorbidities and stroke etiology across the GRS. The Cox proportional hazard model was used to estimate hazard ratios (HRs) and 95% CIs for mortality associated with GRS.

**Results:** The ORs for atrial fibrillation were significantly higher in those at Intermediate GRS [20–80^th^ percentile of GRS; ORs 1.59 (1.25-1.90)] and High GRS [top 20^th^ percentile of GRS; ORs 2.12 (1.69-2.67)] after a full adjustment than in those at Low GRS (bottom 20^th^ percentile of GRS). Regarding stroke etiology, the ORs for cardioembolism were significantly higher in those at Intermediate GRS [ORs 1.31 (1.04-1.61)] and High GRS [ORs 1.44 (1.13-1.89)] than in those at Low GRS. During a median follow-up of 10.0 years, the risk of stroke mortality was significantly higher in those at High GRS [HRs 1.27 (1.04-1.56)] than in those at Low GRS in a fully adjusted model.

**Conclusions:** In Japanese, a higher GRS was significantly associated with atrial fibrillation, cardioembolism, and stroke mortality. Our findings suggest that the GRS may predict the risk of stroke mortality and provide insights into the pathogenesis of stroke.

## Introduction

Stroke is the second-leading cause of death and the primary cause of neurological disability worldwide.^1,2^ Stroke is caused by a complex interplay of environmental and traditional risk factors, including older age, hypertension, diabetes mellitus, dyslipidemia, atrial fibrillation, chronic kidney disease, and smoking.^3^ Besides conventional clinical risk factors, the genetic contribution to the development of stroke is also widely recognized.^4^ Twin and family history studies suggest genetic factors are responsible for some of this unexplained risk for stroke. The heritability estimates were 0.32 for the liability to stroke death and 0.17 for stroke hospitalization or stroke death.^5^

Over the last decades, several genome-wide association studies (GWAS) have identified genetic variants associated with stroke in different ethnic populations.^6–11^ Previous multi-ancestry GWAS of 5,2000 subjects in predominantly European-ancestry groups have identified 32 loci associated with stroke and stroke subtypes (MEGASTROKE study).^11^ Recent work has highlighted the potential of the genetic risk score (GRS) based on the MEGASTROKE study, which can be evaluated as a risk factor for stroke and used to predict incident stroke events in an independent population.^11–15^ The risk of incident stroke was higher in those at high genetic risk than in those at low genetic risk.^12^ The polygenic risk score (PRS) using 3.6 million genetic variants predicts stroke incidents in a population of 12,792 healthy older individuals enrolled in the ASPREE trial (Aspirin in Reducing Events in the Elderly).^13^ In a genetic cohort analysis pooling 51,288 subjects with cardiometabolic disease from five cardiovascular clinical trials, GRS using the set of 32 SNPs derived from the MEGASTROKE study was a strong, independent predictor of ischemic stroke incidence over a median follow-up period of 2.5 years.^14^ Although investigation of genetic risk for stroke has been limited in non-European populations, the Hisayama Study, which involved 3,038 Japanese individuals, reported the PRS for stroke using 350,000 SNPs was significantly associated with stroke incidence during long-term follow-up (median 10.2 years).^15^ Most of the advanced literature on genetic risk for stroke has been reported in general populations regardless of ethnicity; however, solid evidence in the relevant literature has not described the clinical significance of genetic risk for stroke in non-European stroke patients.

To address these limitations, we developed a GRS for stroke from a set of 32 stroke risk loci identified in the MEGASTROKE study in Japanese patients with ischemic stroke. We hypothesized that subsets with a higher genetic risk influence stroke mechanisms and mortality compared to those with a lower genetic risk of ischemic stroke. This cohort study aimed to clarify the association between the GRS score, clinical characteristics, and mortality in stroke patients registered in the BioBank Japan (BBJ) database.

## Methods

### Study participants

The BBJ is a multi-institutional hospital-based registry initially designed to focus on human genetic research.^16,17^ All study participants were Japanese individuals registered in the BBJ project (https://biobankjp.org/). The project aimed to register patients with newly developed diseases (incident cases) as well as those who had been diagnosed and treated before the project started (prevalent cases). The project participants were recruited between June 2003 and March 2018. The biological samples and clinical information were collected and anonymized onsite at the cooperating hospitals. The BBJ 1^st^ cohort consisted of approximately 200,000 patients with 47 common diseases between 2003 and 2007, while the BBJ 2^nd^ cohort included approximately 67,000 patients with 38 diseases between 2013 and 2017. All study participants were diagnosed with one or more of the 51 target diseases, including malignant, cerebral, cardiovascular, respiratory, liver, metabolic, and urologic diseases.^16,17^ The identification of ischemic stroke and comorbidities was based on the physicians’ diagnoses written in the medical records and the questionnaire. If detailed medical record surveys were available, stroke subtypes were determined according to the Trial of ORG 10172 in Acute Stroke Treatment (TOAST) criteria.^18^ For the genetic association analysis, individuals without any history of stroke or intracranial aneurysm were selected as controls (n = 159,610).

### Clinical information

Clinical information, including common clinical variables, disease-specific variables, and laboratory parameters, was collected from each participant upon registration. Among ischemic stroke patients, the following clinical variables were collected: 1) age and sex; 2) systolic and diastolic blood pressure; 3) vascular risk factors, such as hypertension, diabetes, dyslipidemia, and chronic renal failure; 4) atrial fibrillation; 5) congestive heart failure; 6) prior history of ischemic stroke and myocardial infarction; 7) smoking status; 8) alcohol consumption; 9) laboratory parameters; and 10) stroke subtype (large artery atherosclerosis [LA]), cardioembolism [CE], small-vessel occlusion [SVO], and stroke of other & undetermined etiology [O/U]).

### Genotyping, imputation, and quality control

Genomic DNA was extracted from peripheral blood leukocytes using a standard method and genotyped using the Illumina HumanOmniExpress Exome BeadChip array or a combination of the Illumina HumanOmniExpress and HumanExome BeadChips in BBJ1. However, BBJ2 utilized the Illumina Infinium Asian Screening Array for SNP genotyping. We imputed genotypes using the combined reference panel of the 1000 Genomes Project Phase 3 reference panel^19^ and the Japanese in-house reference panel from BBJ using Eagle v2.4.1 and Minimac4 v1.0.2.^20^

Rigorous quality control filters were applied prior to phasing and imputation, including a criterion that excluded variants with sample low call rate <0.98, heterozygous genotype count (HGC) <5, SNP cell rate <0.99, and Hardy-Weinberg equilibrium P< 1.0×10^−6^.

### Calculating GRS and risk categories

We summarized ORs and 95% CIs for 32 stroke risk loci in the MEGASTROKE study and BBJ cohorts (eTable 1). Among these, three SNPs (rs 146390073, rs 12124533, and rs635634) were not assessed in the BBJ cohort. We then constructed a GRS for ischemic stroke using the remaining 29 SNPs. The GRS was calculated following the equation with the use of PLINK v2.0; 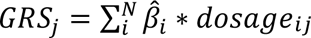; where N is the number of SNPs in the score, 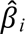 is the effect size of variant *I* and dosage*_ij_* is the number of copies is SNP *i* in the genotype of individual *j*.^21^ Patients were assigned into three categories according to the GRS; “Low GRS” (bottom 20th percentile of GRS), “Intermediate GRS” (20-80th percentile of GRS), and “High GRS” (top 20th percentile of PRS). Regarding the risk categories, we set “Low GRS” as a reference group.

### Statistical analysis

#### Association between GRS and comorbidities

We compared the clinical variables, including age, sex, and comorbidities for stroke, according to the risk categories of the GRS in all patients (n = 19,702). Continuous variables were expressed as medians and interquartile ranges (IQR) in the text and tables. The significance of intergroup differences was assessed using Fisher’s exact test for categorical variables and the Kruskal–Wallis test for continuous variables with Bonferroni correction. Statistical significance was set at P < 0.05. In addition to univariate analysis, we investigated the association between the GRS (continuous and risk categories) and each comorbidity using multivariate logistic analysis. We included sex and age as covariates in the sex- and age-adjusted models (Model 1). The fully adjusted model (Model 2) also included sex, age, comorbidities (hypertension, dyslipidemia, diabetes mellitus, atrial fibrillation, congestive heart failure, and chronic kidney failure), history of ischemic stroke and myocardial infarction, smoking, and alcohol consumption. The logistic regression models were used to estimate odds ratios (ORs) and 95% confidence intervals (95% CIs) for each comorbidity across the GRS (continuous and risk categories).

#### Association between GRS and stroke subtype

Similarly, we also investigated the association between the GRS and stroke subtype in patients with an identified stroke etiology based on the TOAST criteria (n = 6,608). Univariate and multivariate analyses were conducted using the same methods used to evaluate the significant association between the GRS and comorbidities. The significance of intergroup differences was assessed using Fisher’s exact test for categorical variables with Bonferroni correction. Statistical significance was set at P < 0.05. The logistic regression model was used to estimate ORs and 95% confidence intervals (95% CIs) for each stroke subtype across the GRS (continuous and risk categories).

#### Association between GRS and mortality

For the survival analysis, we obtained survival follow-up data based on the cause of death under the ICD-10 code. We assessed Cox proportional hazards models to explore the association between GRS and the risk of mortality (all-cause, stroke, and cardiovascular [CV]) during long-term follow-up (n = 15,468). Models 1 and 2 were similar to those used in the “Association between GRS and comorbidities” subsection. The Cox proportional hazard model was used to estimate hazard ratios (HRs) and 95% CIs for all-cause, stroke, and cardiovascular mortality associated with the GRS (continuous and risk categories).

All statistical analyses were performed using SPSS Software version 25.0 (IBM), GraphPad Prism 10 (GraphPad), and R version 4.0.0 (R Project for Statistical Computing).

#### Standard Protocol Approvals, Registrations, and Patient Consents

Written informed consent was obtained from all participants, and the study was approved by the ethics committees of Nippon Medical School (A-2021-070) and the University of Tokyo (2019-17-0718).

#### Data Availability Statement

All the data used for the analysis are presented in the tables and figures in this article. Data will be shared after obtaining ethical approval if requested by any qualified investigator to replicate the results.

## Results

### Genome-wide significant stroke loci in the BBJ cohort and baseline clinical characteristics

A total of 267,309 individuals with one or more of the 51 target diseases were registered in the BioBank Japan Project between April 2003 and March 2018. Among them, 21,404 individuals with ischemic stroke were identified based on the diagnoses of physicians. After the quality check, 19,702 patients in the BBJ cohort were examined to generate the GRS for ischemic stroke (Table S1).

Table 1 presents the baseline demographic characteristics of the study population. The median age of the participants was 71 years, and 63.1% were male. At the time of registration, blood pressure levels and laboratory parameters were not assessed in any of the subjects. Many patients had traditional comorbidities for stroke, including smoking (53.2%), alcohol (51.4%), previous ischemic stroke (43.6%), hypertension (37.0%), dyslipidemia (18.2%), and diabetes (15.7%). A small proportion of the cohort had myocardial infarction (4.6%), atrial fibrillation (4.4%), congestive heart failure (3.0%), or chronic kidney failure (2.1%). The stroke subtype was identified in 6,608 patients based on the TOAST criteria (Figure S1: LAA; n=1276, CE; n=752, SVO; n=3687, O/U; n=893).

**Table 1:** Baseline demographics.

|  | All (n=19,702) |
| --- | --- |
| Men, n (%) | 12,441 (63.1) |
| Age, year-old; median (IQR) | 71 (64-77) |
| Blood pressure level at registration, mmHg; median (IQR) [n=16,672]* |  |
| Systolic blood pressure | 134 (124-145) |
| Diastolic blood pressure | 80 (70-84) |
| Comorbidities, n (%) |  |
| Hypertension | 7,297 (37.0) |
| Dyslipidemia | 3,586 (18.2) |
| Diabetes mellitus | 3,090 (15.7) |
| Atrial fibrillation | 866 (4.4) |
| Congestive heart failure | 588 (3.0) |
| Chronic kidney failure | 409 (2.1) |
| Previous ischemic stroke | 8,598 (43.6) |
| Myocardial infarction | 902 (4.6) |
| Smoking | 10,474 (53.2) |
| Alcohol | 10,123 (51.4) |
| Laboratory parameters at registration |  |
| WBC, $\times 100/\mu\text{l}$; median (IQR) [n=18,288]* | 63 [52-77] |
| RBC, $\times 10000/\mu\text{l}$; median (IQR) [n=18,706]* | 446 [410-480] |
| Hb, g/dl; median (IQR) [n=18,708]* | 13.9 [12.8-15.0] |
| Plt, $\times 10000/\mu\text{l}$; median (IQR) [n=17,582]* | 21.8 [18.0-25.9] |
| AST, IU/L; median (IQR) [n=18,084]* | 22 [18-27] |
| ALT, IU/L; median (IQR) [n=17,872]* | 19 [14-27] |
| BUN, md/dL; median (IQR) [n=16,442]* | 15.9 [13.0-19.6] |
| Cr, mg/dl; median (IQR) [n=17,701]* | 0.80 [0.69-1.00] |
| eGFR, ml/min/1.73m<sup>2</sup>; median (IQR) [n=17,701]* | 65.4 [54.0-77.4] |
| TP, mg/dL; median (IQR) [n=10,900]* | 7.1 [6.7-7.4] |
| Alb, mg/dL; median (IQR) [n=8,973]* | 4.2 [3.9-4.4] |
| TC, mg/dL; median (IQR) [n=14,354]* | 197 [173-221] |
| LDL-C, mg/dL; median (IQR) [n=4,292]* | 112 [93-135] |
| HDL-C, mg/dL; median (IQR) [n=9,605]* | 51 [43-62] |
| TG, mg/dL; median (IQR) [n=13,259]* | 118 [85-170] |
| Glucose, mg/dl; median (IQR) [n=13,956]* | 111 [97-142] |
| HbA1c, %; median (IQR) [n=4,959]* | 5.8 [5.4-6.7] |
| Stroke subtype, n (%) |  |
| Cardioembolism | 752 (3.8) |
| Large artery atherosclerosis | 1,276 (6.5) |
| Small vessel occlusion | 3,687 (18.7) |
| Others/Undetermined | 893 (4.5) |
| Etiology N/A** | 13,094 (66.4) |
| Genetic risk score; median (IQR) | 2.12 (1.88-2.35) |

IQR; interquartile range, WBC; white blood cell, RBC; red blood cell, Hb; hemoglobin, Plt; platelet, AST; aspartate aminotransferase, ALT; alanine aminotransferase, BUN; blood urea nitrogen, Cr; creatinine, eGFR; estimated glomerular filtration rate, TP; total protein, Alb; albumin, TC; total cholesterol, LDL-C; low-density lipoprotein cholesterol, HDL-C; high-density lipoprotein cholesterol, TG; triglyceride, HbA1c; hemoglobin A1c, N/A; not available.

### Association between GRS and comorbidities

Baseline characteristics by GRS risk category are shown in Table S2. Subjects at High GRS were more likely to be younger and more male-dominant than those at Low GRS. The prevalence of atrial fibrillation was significantly higher among patients with Intermediate and High GRS than those at Low GRS. The correlation between the GRS, blood pressure, and laboratory parameters is shown in Figure S2. We investigated the association between GRS (both continuous level and risk categories) and each comorbidity using multivariate logistic analysis (Table 2). The ORs (95%CIs) of atrial fibrillation in those at Intermediate and High GRS were significantly higher in the fully adjusted model than those at Low GRS. Similarly, higher GRS (continuous level) was significantly associated with atrial fibrillation in the fully adjusted model (Table 2 and Figure 1). A significant association was also observed between GRS and hypertension in the fully adjusted model (Table 2 and Figure 1).

**Figure 1.**
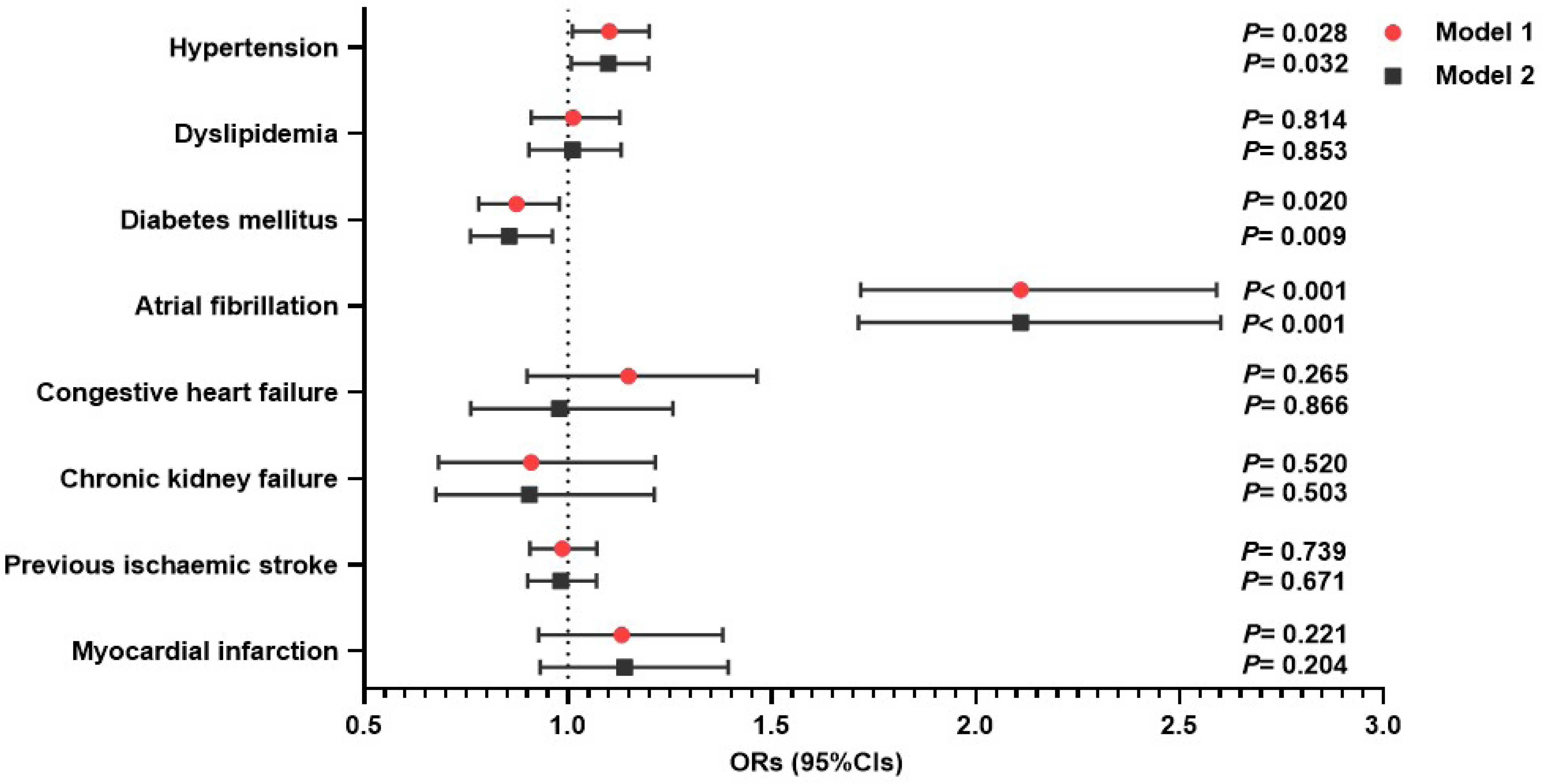
Impact of the continuous genetic risk score (GRS) for stroke and comorbidities (n=19702). The data are presented as estimated odds ratios (ORs) and 95% confidence intervals (CIs) for increased GRS for stroke. Statistical significance was set at P < 0.05. **Model 1:** Adjusted for age and sex. **Model 2:** Adjusted for Model 1 + other stroke comorbidities.

**Table 2.**
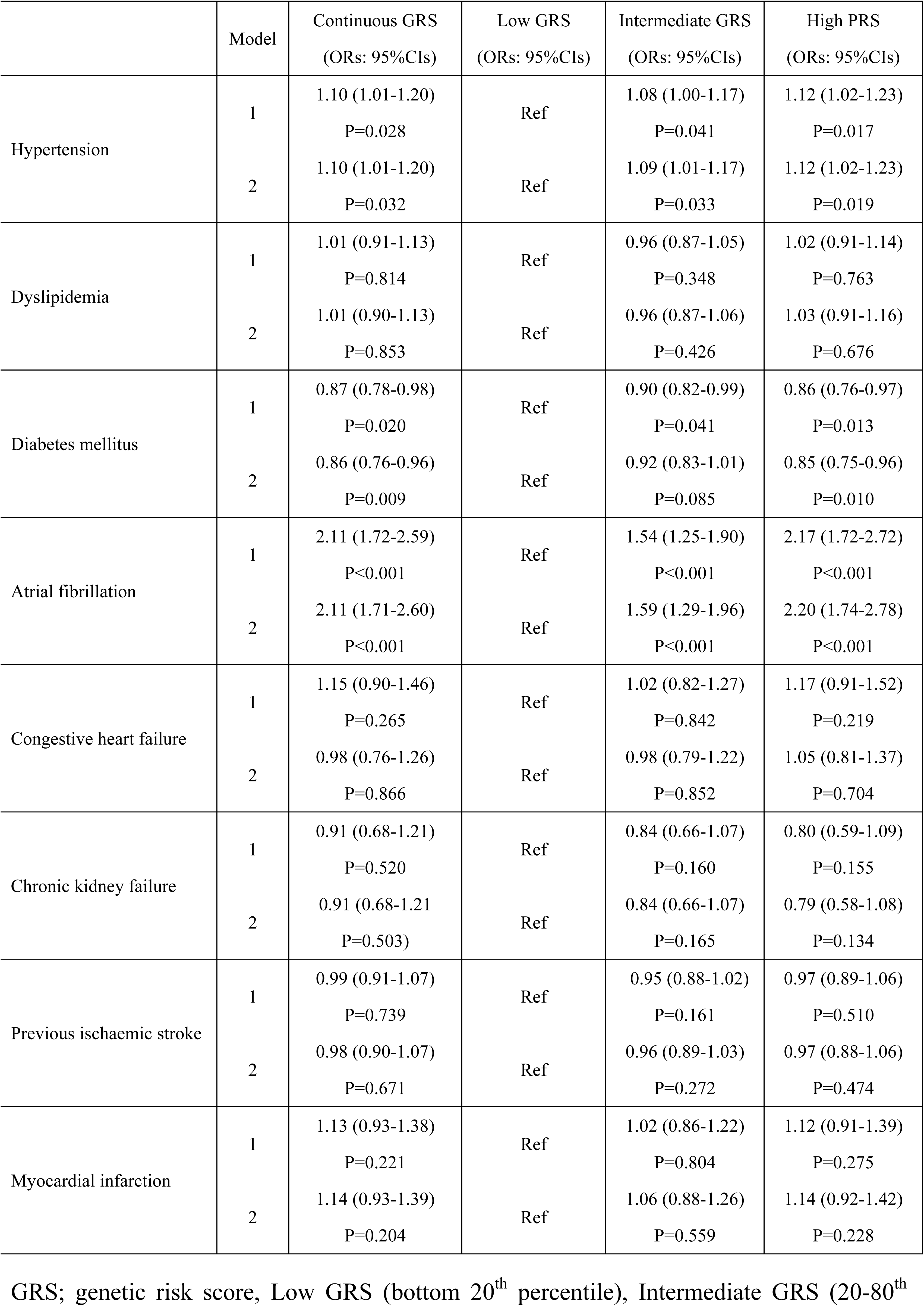
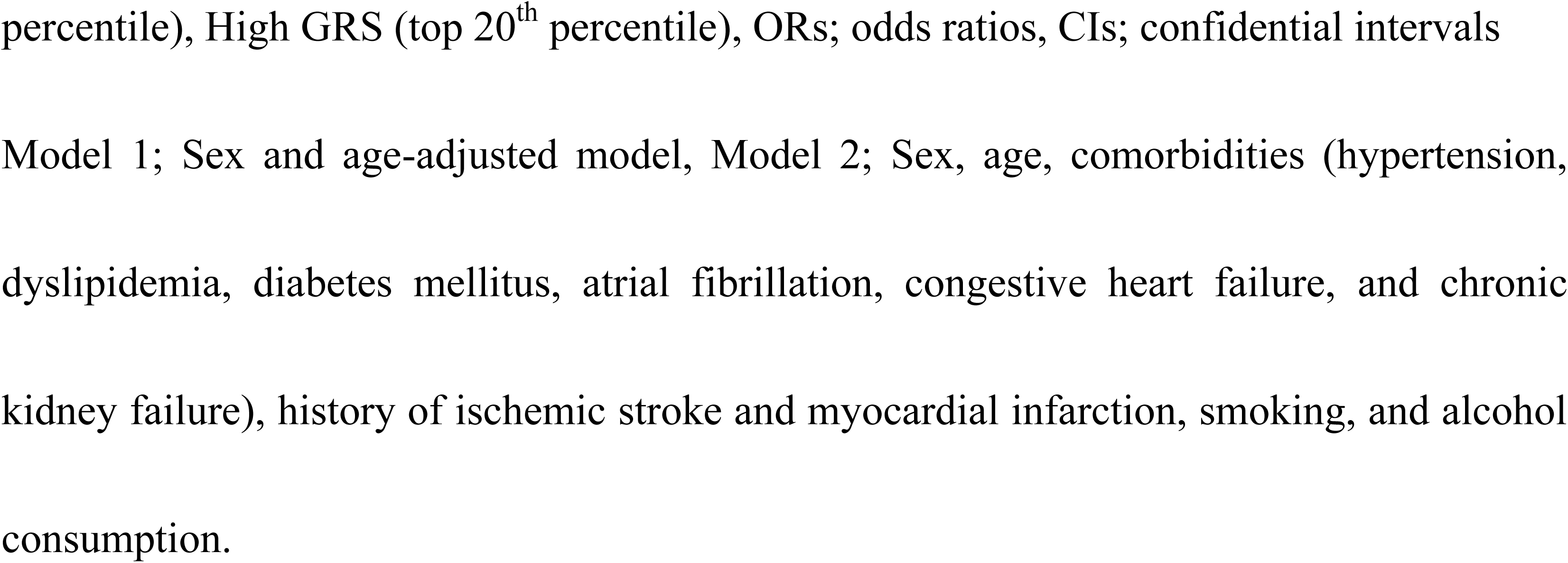
Association between genetic risk score (GRS) and comorbidities by multivariate logistic analysis.

|  | Model | Continuous GRS<br>(ORs: 95% CIs) | Low GRS<br>(ORs: 95% CIs) | Intermediate GRS<br>(ORs: 95% CIs) | High PRS<br>(ORs: 95% CIs) |
| --- | --- | --- | --- | --- | --- |
| Hypertension | 1 | 1.10 (1.01-1.20)<br>P=0.028 | Ref | 1.08 (1.00-1.17)<br>P=0.041 | 1.12 (1.02-1.23)<br>P=0.017 |
|  | 2 | 1.10 (1.01-1.20)<br>P=0.032 | Ref | 1.09 (1.01-1.17)<br>P=0.033 | 1.12 (1.02-1.23)<br>P=0.019 |
| Dyslipidemia | 1 | 1.01 (0.91-1.13)<br>P=0.814 | Ref | 0.96 (0.87-1.05)<br>P=0.348 | 1.02 (0.91-1.14)<br>P=0.763 |
|  | 2 | 1.01 (0.90-1.13)<br>P=0.853 | Ref | 0.96 (0.87-1.06)<br>P=0.426 | 1.03 (0.91-1.16)<br>P=0.676 |
| Diabetes mellitus | 1 | 0.87 (0.78-0.98)<br>P=0.020 | Ref | 0.90 (0.82-0.99)<br>P=0.041 | 0.86 (0.76-0.97)<br>P=0.013 |
|  | 2 | 0.86 (0.76-0.96)<br>P=0.009 | Ref | 0.92 (0.83-1.01)<br>P=0.085 | 0.85 (0.75-0.96)<br>P=0.010 |
| Atrial fibrillation | 1 | 2.11 (1.72-2.59)<br>P<0.001 | Ref | 1.54 (1.25-1.90)<br>P<0.001 | 2.17 (1.72-2.72)<br>P<0.001 |
|  | 2 | 2.11 (1.71-2.60)<br>P<0.001 | Ref | 1.59 (1.29-1.96)<br>P<0.001 | 2.20 (1.74-2.78)<br>P<0.001 |
| Congestive heart failure | 1 | 1.15 (0.90-1.46)<br>P=0.265 | Ref | 1.02 (0.82-1.27)<br>P=0.842 | 1.17 (0.91-1.52)<br>P=0.219 |
|  | 2 | 0.98 (0.76-1.26)<br>P=0.866 | Ref | 0.98 (0.79-1.22)<br>P=0.852 | 1.05 (0.81-1.37)<br>P=0.704 |
| Chronic kidney failure | 1 | 0.91 (0.68-1.21)<br>P=0.520 | Ref | 0.84 (0.66-1.07)<br>P=0.160 | 0.80 (0.59-1.09)<br>P=0.155 |
|  | 2 | 0.91 (0.68-1.21)<br>P=0.503 | Ref | 0.84 (0.66-1.07)<br>P=0.165 | 0.79 (0.58-1.08)<br>P=0.134 |
| Previous ischaemic stroke | 1 | 0.99 (0.91-1.07)<br>P=0.739 | Ref | 0.95 (0.88-1.02)<br>P=0.161 | 0.97 (0.89-1.06)<br>P=0.510 |
|  | 2 | 0.98 (0.90-1.07)<br>P=0.671 | Ref | 0.96 (0.89-1.03)<br>P=0.272 | 0.97 (0.88-1.06)<br>P=0.474 |
| Myocardial infarction | 1 | 1.13 (0.93-1.38)<br>P=0.221 | Ref | 1.02 (0.86-1.22)<br>P=0.804 | 1.12 (0.91-1.39)<br>P=0.275 |
|  | 2 | 1.14 (0.93-1.39)<br>P=0.204 | Ref | 1.06 (0.88-1.26)<br>P=0.559 | 1.14 (0.92-1.42)<br>P=0.228 |

GRS; genetic risk score, Low GRS (bottom 20<sup>th</sup> percentile), Intermediate GRS (20-80<sup>th</sup>
percentile), High GRS (top 20<sup>th</sup> percentile), ORs; odds ratios, CIs; confidential intervals
Model 1; Sex and age-adjusted model, Model 2; Sex, age, comorbidities (hypertension, dyslipidemia, diabetes mellitus, atrial fibrillation, congestive heart failure, and chronic kidney failure), history of ischemic stroke and myocardial infarction, smoking, and alcohol consumption.

### Association between GRS and stroke subtype

The prevalence of stroke subtypes based on the TOAST criteria by GRS category is summarized in Table S3. Subjects in Intermediate and High GRS were more likely to have CE than those at Low GRS. Conversely, those with Lower and Intermediate GRS were more likely to have SVO than those at High GRS. We investigated the association between GRS (both continuous level and risk categories) and stroke subtype using multivariate logistic analysis (Table 3). The ORs (95%CIs) for CE were significantly higher in those at Intermediate GRS and High GRS in the fully adjusted model (Table 3). Higher GRS (continuous level) was significantly associated with CE in the fully adjusted model (Table 3 and Figure 2). However, a lower GRS was significantly associated with SVO in the fully adjusted model (Table 3 and Figure 2).

**Figure 2.**
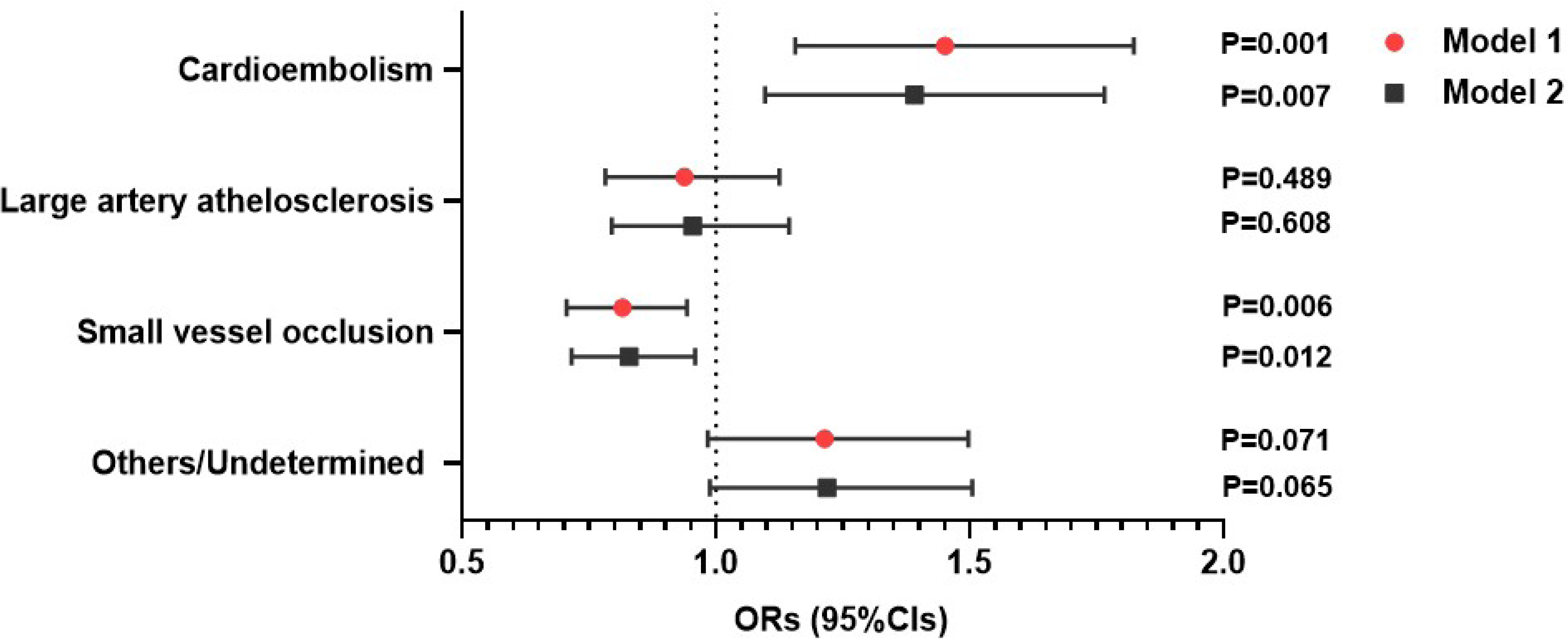
Impact of continuous genetic risk score (GRS) for stroke and stroke subtypes based on TOAST criteria (n = 6608). The data are presented as estimated odds ratios (ORs) and 95% confidence intervals (CIs) for increased GRS for stroke. Statistical significance was set at P < 0.05. **Model 1:** Adjusted for age and sex. **Model 2:** Adjusted for Model 1 + other stroke comorbidities.

**Table 3.** Association between genetic risk score (GRS) and stroke etiology by multivariate logistic analysis.

|  | Model | Continuous GRS<br>(ORs: 95% CIs) | Low GRS<br>(ORs: 95% CIs) | Intermediate GRS<br>(ORs: 95% CIs) | High GRS<br>(ORs: 95% CIs) |
| --- | --- | --- | --- | --- | --- |
| Cardioembolism | 1 | 1.45 (1.16-1.82)<br>P=0.001 | Ref | 1.37 (1.11-1.70)<br>P=0.004 | 1.48 (1.15-1.90)<br>P=0.002 |
|  | 2 | 1.39 (1.10-1.77)<br>P=0.007 | Ref | 1.31 (1.05-1.64)<br>P=0.016 | 1.44 (1.11-1.87)<br>P=0.006 |
| Large artery atherosclerosis | 1 | 0.94 (0.78-1.13)<br>P=0.489 | Ref | 0.89 (0.76-1.04)<br>P=0.143 | 0.96 (0.80-1.16)<br>P=0.692 |
|  | 2 | 0.95 (0.80-1.14)<br>P=0.608 | Ref | 0.90 (0.77-1.05)<br>P=0.184 | 0.97 (0.80-1.18)<br>P=0.775 |
| Small vessel occlusion | 1 | 0.82 (0.71-0.94)<br>P=0.006 | Ref | 0.95 (0.83-1.07)<br>P=0.380 | 0.83 (0.71-0.97)<br>P=0.019 |
|  | 2 | 0.83 (0.72-0.96)<br>P=0.012 | Ref | 0.96 (0.84-1.09)<br>P=0.520 | 0.84 (0.72-0.99)<br>P=0.032 |
| Others/Undetermined | 1 | 1.21 (0.98-1.50)<br>P=0.071 | Ref | 1.03 (0.86-1.24)<br>P=0.760 | 1.13 (0.90-1.41)<br>P=0.292 |
|  | 2 | 1.22 (0.99-1.51)<br>P=0.065 | Ref | 1.03 (0.86-1.24)<br>P=0.732 | 1.13 (0.91-1.41)<br>P=0.278 |

GRS; genetic risk score, Low GRS (bottom 20th percentile), Intermediate GRS (20-80th percentile), High GRS (top 20th percentile), ORs; odds ratios, CIs; confidential intervals.
Model 1; Sex and age-adjusted model, Model 2; Sex, age, comorbidities (hypertension, dyslipidemia, diabetes mellitus, atrial fibrillation, congestive heart failure, and chronic kidney failure), history of ischemic stroke and myocardial infarction, smoking, and alcohol consumption.

### Association between GRS and mortality

During the median follow-up of 10.0 years, 15,468 patients were included in the outcome analysis (Figure S1). The number of events for mortality was as follows (Table 4): all-cause (n=6,253, 31.7%), stroke (n=949, 4.8%), and cardiovascular (n=1,167, 5.9%). Kaplan–Meier estimates of cumulative mortality rate were higher in those at High GRS than those at Low GRS stroke-related mortality for stroke and CV mortality by Cox proportional hazard analysis (High GRS: HRs 1.27 [1.04-1.56], P=0.018 for stroke mortality; High GRS: HRs 1.27 [1.06-1.53], P=0.009 for CV mortality) (Table 4 and Figure 3). These significant associations were observed through continuous GRS level (HRs 1.30 [1.07-1.56], P=0.007 for stroke mortality: HRs 1.26 [1.06-1.50], P=0.007 for CV mortality) (Table 4 and Figure 3). However, no significant association was observed between the GRS and all-cause mortality using the Cox proportional hazard analysis (Table 4 and Figure 3).

**Figure 3.**
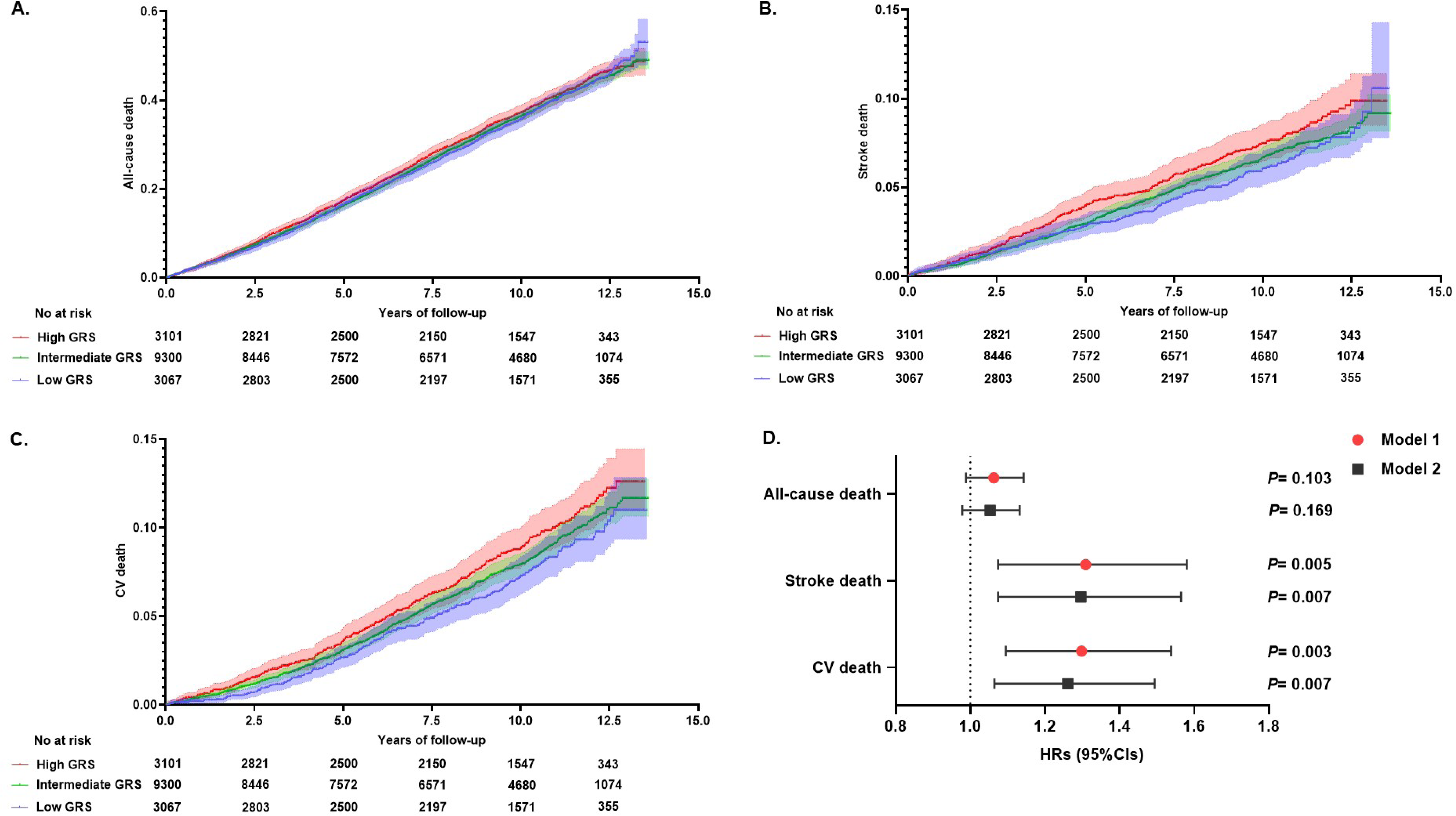
Impact of genetic risk score (GRS) for stroke on long-term mortality in Cox proportional hazard analysis. Kaplan-Meier estimates of cumulative events from **(A)** all-cause, **(B)** stroke, and **(C)** cardiovascular (CV) are shown with a band of 95% CIs. Individuals were classified as having high GRS (red), intermediate GRS (green), or low GRS (blue). **(D)** Effect of continuous GRS for stroke and mortality (all-cause, stroke, and CV). Data are presented as estimated hazard ratios (HRs) and 95% confidence intervals (CIs) for increased GRS for stroke. Statistical significance was set at P < 0.05. **Model 1:** Adjusted for age and sex. **Model 2:** Adjusted for Model 1 + other stroke comorbidities.

**Table 4.** Association between genetic risk score (GRS) and mortality by Cox proportional hazard analysis (n=15,468).

|  | No of events (%) | Model | GRS continuous<br>(HRs: 95% CIs) | Low GRS<br>(HRs: 95% CIs) | Intermediate GRS<br>(HRs: 95% CIs) | High GRS<br>(HRs: 95% CIs) |
| --- | --- | --- | --- | --- | --- | --- |
| All-cause mortality | 6,253 (31.7%) | 1 | 1.06 (0.99-1.14)<br>P=0.103 | Ref | 1.03 (0.96-1.10)<br>P=0.395 | 1.08 (0.99-1.17)<br>P=0.052 |
|  |  | 2 | 1.05 (0.98-1.13)<br>P=0.169 | Ref | 1.03 (0.97-1.10)<br>P=0.348 | 1.08 (0.99-1.17)<br>P=0.063 |
| Stroke mortality | 949 (4.8%) | 1 | 1.31 (1.07-1.58)<br>P=0.005 | Ref | 1.09 (0.92-1.29)<br>P=0.322 | 1.28 (1.05-1.56)<br>P=0.016 |
|  |  | 2 | 1.30 (1.07-1.56)<br>P=0.007 | Ref | 1.09 (0.92-1.29)<br>P=0.323 | 1.27 (1.04-1.56)<br>P=0.018 |
| CV mortality | 1,167 (5.9%) | 1 | 1.30 (1.10-1.54)<br>P=0.003 | Ref | 1.14 (0.97-1.32)<br>P=0.107 | 1.30 (1.09-1.56)<br>P=0.004 |
|  |  | 2 | 1.26 (1.06-1.50)<br>P=0.007 | Ref | 1.13 (0.97-1.32)<br>P=0.120 | 1.27 (1.06-1.53)<br>P=0.009 |

GRS; genetic risk score, Low GRS (bottom 20<sup>th</sup> percentile), Intermediate GRS (20-80<sup>th</sup> percentile), High GRS (top 20<sup>th</sup> percentile), HRs; hazard ratios, CIs; confidential intervals, CV; cardiovascular.
Model 1; Sex and age-adjusted model, Model 2; Sex, age, comorbidities (hypertension, dyslipidemia, diabetes mellitus, atrial fibrillation, congestive heart failure, and chronic kidney failure), history of ischemic stroke and myocardial infarction, smoking, and alcohol consumption.

## Discussion

We constructed a GRS for stroke using 29 SNPs from a set of 32 stroke-related loci in Japanese stroke patients. The previous MEGASTROKE study has predominantly involved Western populations,^11^ and the impact of stroke risk loci has not been thoroughly examined in Japanese patients with ischemic stroke. The genetic loci rs7610618 and rs10820405, which are associated with LAA in the European population,^11^ were not statistically significant in the BBJ cohort. In a cohort study of 30,6473 UK Biobank, 90 SNPs were used to assess the significance of stroke incidence risk during a median follow-up of 7.1 years.^12^ In the ASPREE trial, PRS was calculated from 3.6 million SNPs in 12,792 elderly individuals to investigate its association with stroke incidence risk.^13^ Among 51,288 subjects enrolled in five clinical trials across the spectrum of cardiometabolic diseases, GRS using a set of SNPs based on the MEGASTROKE study was a strong, independent predictor of ischemic stroke incidence.^14^ In the Hisayama Study, which involved 3,038 Japanese individuals, the PRS of stroke was calculated from 350,000 SNPs.^15^ The high PRS group exhibited HRs of 1.63 for stroke incidence compared with the low PRS group. However, this cohort developed a small number of stroke incidences (91 cases) during follow-up (median 10.2 years).^15^ Additionally, the recent GIGASTROKE study identified 89 SNPs as stroke-related genes and examined the association between integrative PRS and stroke incidence.^22^ Therefore, selecting optimal SNPs to create GRS varies depending on racial differences, sample size, and target population (healthy individuals and patients with cardiovascular diseases or stroke).

We found a strong association between the GRS and atrial fibrillation in Japanese patients with ischemic stroke. A high genetic risk for stroke increases the incidence of stroke development during long-term follow-up in the general population, healthy older individuals, and patients with cardiovascular issues.^11–15^ However, clear evidence has not indicated whether the genetic risk for stroke is associated with vascular risk factors in non-European patients with ischemic stroke. In the MEGASTROKE study, approximately half of the identified loci shared genetic variation with related vascular traits, including blood pressure, atrial fibrillation, and lipid levels.^11^ In the BBJ cohort, the ORs of loci for atrial fibrillation (rs13143308; ORs 1.41, rs 12932445; ORs 1.21) were higher than the ORs of other loci for blood pressure (rs 880315; ORs 1.06, rs 1689638; ORs 1.05, rs4932370; ORs 1.07, rs35436, ORs 1.08) and lipid disorders (rs8103309; ORs 1.06). Recent cross-ancestry GWAS of atrial fibrillation identified 35 new susceptibility loci using data from BBJ and European cohorts (77,690 cases, 1,167,040 controls).^23^ Notably, the PRS for atrial fibrillation predicted increased risks of CV and stroke mortalities and segregated individuals with cardioembolic stroke in undiagnosed atrial fibrillation patients.^23^ Moreover, Ebara et al. reported that GRS using eight risk loci for atrial fibrillation was associated with the risk of ischemic stroke in patients with atrial fibrillation in the BBJ cohort.^24^ Therefore, the genetics of atrial fibrillation may play a clinically important role in Japanese patients with ischemic stroke.

Among patients with an identified stroke etiology based on the TOAST criteria, a higher GRS was also associated with CE and atrial fibrillation. In contrast, we found a significant association between lower GRS and SVO. A previous Japanese cohort study using two independent data sets (Kyusyu U data set and JPJM data set), showed that the ORs of the top PRS quintiles were significantly higher than those of the lowest PRS quintiles when compared with the control group in CE (134 cases, 134 matched controls), LAA (360 cases, 360 matched controls), and SVO (486 cases, 486 matched controls).^25^ In the ASPREE trial, continuous GRS indicated a significant predictor for the risk of large vessel and cardioembolic stroke but not for small vessel stroke among 12,792 healthy older individuals over 5 years.^13^ Our findings suggest that a higher GRS for stroke may be helpful for identifying high-risk sources of CE, including atrial fibrillation, in Japanese patients with ischemic stroke. Identifying the underlying stroke etiology is important because its pathophysiology has consequences for acute treatment and secondary stroke prevention. Traditionally, an ischemic stroke with an unclear etiology based on the TOAST criteria was classified as cryptogenic stroke.^26^ In 2014, the concept of embolic stroke of undetermined source (ESUS) was developed based on previous observations that patients with non-lacunar cryptogenic ischemic stroke were likely to have embolic stroke mechanisms.^26,27^ Occult paroxysmal atrial fibrillation has been considered an important cause of ESUS.^28,29^ Atrial fibrillation was detected in 5.1% of patients in the in-hospital setting, and 8.9%, 12.4%, and 30.0% at 6, 12, and 36 months, respectively, after stroke with an insertable cardiac monitor (ICM).^28,29^ To date, no detailed investigations have been conducted regarding GRS and the diagnostic work of ESUS with ICM, further research is needed.

In the present study, we confirmed a significant association between the GRS for stroke and the risk of stroke and CV mortality during a long-term follow-up. Previous cohort studies have shown a significant association between a higher genetic risk for stroke and the incidence of future stroke in the general population.^11,12,22^ However, solid evidence in the relevant literature has not described the clinical relevance of the GRS for stroke to survival prediction in patients with stroke. A previous GWAS for atrial fibrillation and coronary artery disease showed that a higher PRS for these diseases significantly increased the risk of CV mortality in the BBJ cohort.^23,30^ Therefore, our findings were strengthened by the large population-based cohort design with a long-term follow-up to explore the association between the GRS for stroke and stroke mortality as well as cardiovascular disease in Japanese individuals.

This study had several limitations. First, the cause of mortality was identified based on the ICD-10 codes. Clinical information in the BBJ cohort may not include as many clinical details as individual hospital records regarding survival analysis. Second, only one-third of the participants were diagnosed with stroke subtypes based on the TOAST criteria. Moreover, most patients with stroke in the BBJ cohort included those with mild disabilities or in a stable chronic stage. The proportion of SVO accounted for over half (56%), while LAA was at 19% and CE was at 11% in this study. The Japanese stroke data, a hospital-based multicenter acute stroke registration database (n=10392), showed that the proportion of stroke subtypes based on the TOAST criteria were as follows: LAA, 30%; CE at 27%; and SVO, 22%.^31^ The difference in baseline characteristics, including risk factors, stroke subtypes, and neurological severity, should be taken into account when investigating the clinical significance of GRS in patients with ischemic stroke. Third, the brain magnetic resonance imaging (MRI) findings were not assessed in the BBJ cohort. Regarding the association between brain MRI findings and GWAS in the general cohort, white matter lesions (WML),^32^ cerebral microbleeds (CMBs),^33^ and perivascular spaces (PVS),^34^ indicating imaging markers of cerebral small vessel disease, were assessed in the general cohorts. However, it remains unclear whether there exists an association between genetic risk for stroke and other MRI findings, such as acute ischemic volume on diffusion-weighted imaging (DWI) as well as large vessel involvement due to atherosclerotic changes or cardiac embolism. Finally, to confirm an association between the GRS, clinical characteristics, and mortality in a non-European population, our study population included only Japanese subjects from the BBJ cohort. Recent cross-ancestry GWAS meta-analyses of 110,182 stroke patients (GIGASTROKE study) identified 89 independent stroke risk loci.^22^ Higher GIGASTROKE GRS was significantly associated with increased risk of stroke in the East Asian cohort (1,312 participants of whom 27 developed an incident stroke over a 3-year follow-up; HRs =1.49, 95% CIs=1.00–2.21, P = 0.048), whereas the MEGASTROKE GRS was not associated with incident stroke (HRs=0.82, 95% CIs=0.55-1.23, P=0.34).^22^ Further studies, including the GIGASTROKE GRS, brain MRI findings, and clinical outcomes, are needed to clarify the clinical significance of genetic risk in patients with ischemic stroke.

This large cohort study demonstrated an association between the GRS for stroke, clinical characteristics, and mortality in Japanese patients with ischemic stroke. The GRS for stroke was significantly associated with atrial fibrillation, CE, and stroke mortality. Our findings suggest that the GRS for stroke may provide insights into the pathogenesis of stroke and predict the risk of stroke mortality.

## Acknowledgment

We would like to thank all the participants for their willingness and time devoted to this study and extend our appreciation to the BBJ project team for their assistance in data collection.

